# Antibody Responses in Elderly Residential Care Persons following COVID-19 mRNA Vaccination

**DOI:** 10.1101/2021.04.07.21254925

**Authors:** David A. Nace, Kevin E. Kip, Octavia M. Peck Palmer, Michael R. Shurin, Katie Mulvey, Melissa Crandall, April L. Kane, Amy Lukanski, Paula L. Kip, Alan L. Wells

**Affiliations:** Division of Geriatric Medicine, School of Medicine, University of Pittsburgh, Pittsburgh, PA; Clinical Analytics, University of Pittsburgh Medical Center, Pittsburgh, PA, USA; Department of Pathology, University of Pittsburgh, Pittsburgh, PA, USA; Clinical Laboratory, University of Pittsburgh Medical Center, Pittsburgh, PA, USA; Senior Services, University of Pittsburgh Medical Center, Pittsburgh, PA, USA; Wolff Center, University of Pittsburgh Medical Center, Pittsburgh, PA, USA

**Keywords:** COVID-19, SARS-CoV-2, mRNA vaccine, assisted living, independent living, personal care, reopening, quality improvement

## Abstract

**Objective:** COVID-19 disproportionately impacts older adults residing at long-term care facilities. Data regarding antibody response to COVID-19 vaccines in this population is limited. Our objective was to quantify the presence and magnitude of antibody response in older, vaccinated residents at assisted living, personal care, and independent living facilities.

**Design:** A cross-sectional quality improvement study was conducted March 15 – April 1, 2021 in the Pittsburgh region.

**Setting and Population:** Participants were volunteers at assisted living, personal care, and independent living facilities, who received mRNA COVID-19 vaccine. Conditions that obviate immune responses were exclusionary criteria.

**Methods:** Sera were collected to measure IgG anti-SARS-CoV-2 antibody level with reflex to total anti-SARS-CoV-2 immunoglobulin levels. Descriptive statistics, Pearson correlation coefficients, and multiple linear regression analysis were performed to evaluate relationships between factors potentially associated with antibody levels.

**Results:** All participants (N=70) had received two rounds of vaccination for COVID-19 and were found to have antibodies to SARS-CoV-2. There was wide variation in relative levels of antibodies as determined by extinction coefficients. Antibody levels trended lower in male sex, advanced age, steroid medications, and longer length of time from vaccination.

**Conclusions and Implications:** Higher functioning long-term care residents mounted detectable antibody responses when vaccinated with COVID-19 mRNA-based vaccines. This study provides preliminary information on level of population risk of assisted living, personal care, and independent living residents which can inform reopening strategies. Data suggests some degree of immunity is present during the immediate period following vaccination. However, protective effects of such vaccination programs remain to be determined in larger studies. Clinical protection is afforded not just by pre-formed antibody levels, but by ongoing adaptive immunity, which is known to be decreased in older individuals. Thus, the implications of these levels of antibodies in preventing COVID-19 disease must be determined by clinical follow-up.

## INTRODUCTION

COVID-19 disproportionately impacts older adults and frail individuals residing in long-term care facilities. As of March 2021, there are over 1.4 million cases of COVID-19 in U.S. nursing homes. In addition, over 175,000 COVID-19 related deaths have occurred, representing 34% of all U.S. COVID-19 deaths.^1^ Advanced-age, high rates of frailty and comorbid conditions along with close physical contact between residents and staff facilitate spread of the virus in these settings. Visitor restrictions, curtailing of community dining, and other social activities have been crucial to limiting spread of the virus. Between December 2020 and February 2021, the number of nursing home cases decreased by 80% and deaths by 65%, due in part to COVID-19 vaccinations.^2^ Given the reductions in cases and severity, residents and families are now calling for reopening of long-term care facilities to reduce the negative impacts of social isolation on residents. The Centers for Medicare and Medicaid Services released guidance for reopening of nursing homes on March 10, 2021,^3^ but so far, no consensus exists around reopening strategies for independent living, personal care, and assisted living facilities.

While current COVID-19 vaccines appear to be effective in reducing severe illness, breakthrough cases do occur including asymptomatic infections. Age and frailty status are linked to reduced vaccine response for other vaccines. Information regarding antibody response to COVID-19 vaccines is limited. As part of an effort to assess level of risk in reopening strategies, the Society for Post-Acute and Long-Term Care Medicine (AMDA), is recommending a measured, stepwise approach to resuming visitation and group activities in post-acute and long-term care settings while acknowledging gaps in clinical knowledge about COVID-19.^4^ While recommendations regarding reopening have been published,^5-7^ these focus on the process for reopening and not risk assessment of the resident population. Antibody measurement may help inform level of risk, particularly if significant numbers of individuals fail to demonstrate antibody response. Therefore, the objective of this study was to quantify the presence and magnitude of antibody response in older, vaccinated adults residing in assisted living, personal care, and independent living facilities, including those with and without prior COVID-19 infection.

## METHODS

### Setting and Population

A cross-sectional quality improvement study was conducted March 15 – April 1, 2021 at University of Pittsburgh Medical Center (UPMC) Senior Communities assisted living, personal care, and independent living facilities in the Pittsburgh metropolitan region. Participants were selected from volunteers at UPMC Senior Communities to determine antibody responses in the elderly. Participant eligibility criteria were residents who have received one or more doses of a COVID-19 vaccine. Conditions that obviate immune responses were exclusionary criteria; these were hematologic malignancies, solid organ transplants, active chemotherapy, and those that require specific immunosuppressive therapies. Individuals receiving steroids at doses equivalent to less than 20 mg of prednisone daily or for less than ten-days duration were not excluded. This project underwent review and was granted ethical approval as a quality improvement study by the UPMC Quality Improvement Review Committee (Project ID: 3250), the ethics, regulatory, and legal oversight body for protecting patient/participant rights, confidentiality, consent (including waiver of consent), and the analysis and dissemination of deidentified data within the UPMC system.

### Data Collection

Study data were collected and managed using the Research Electronic Data Capture (REDCap) hosted at UPMC.^8^ REDCap is a secure, web-based software platform designed to support data capture for research and quality improvement studies, providing 1) an intuitive interface for validated data capture; 2) audit trails for tracking data manipulation and export procedures; 3) automated export procedures for seamless data downloads to common statistical packages; and 4) procedures for data integration and interoperability with external sources.^9^ Data was collected on vaccination status (number of doses, dates, and type of vaccine), medical conditions, and current medications. Level of frailty was assessed in participants using self-reported activities of daily living and instrumental activities of daily living measures.^10-12^

### Study Outcomes

To quantify the presence and magnitude of antibody response in this population, sera were collected from each participant to measure IgG anti-SARS-CoV-2 antibody level with reflex to total anti-SARS-CoV-2 immunoglobulin levels. SARS-CoV-2 antibody assays were performed in the UPMC Clinical Laboratories at the Clinical Laboratory Building in Pittsburgh, PA. These are CLIA-88 accredited laboratories for clinical testing. The specimens were initially assessed using the Beckman Coulter SARS-CoV-2 IgG Access assay (AU5800 analyzer, Brea, CA, USA), and then confirmed orthogonally using the Siemens Healthineers SARS-CoV-2 Total assay (ADIVA Centaur XP analyzer, Munich, Germany; Siemens-C).^13,14^ The Beckman Coulter assay uses S1 Spike antigens as capture and anti-IgG as reporter; the Siemens uses S1 Spike antigens as both capture and reporter and thus IgM antibodies are detected and at a higher molar ‘index value’ than IgG antibodies. Both assays were run according to the manufacturer’s instructions. Both assays use units that are generated by comparison to an internal calibrator or standard, when referring to assay results collectively we refer to these units as ‘index values’ for simplicity; both use an index of >1.0 for positivity.

### Statistical Methods

Descriptive statistics for baseline population characteristics were calculated as means, standard deviations, and frequencies. Pearson correlation coefficients were calculated between age and antibody level, and between days since vaccination and antibody level. We also performed multiple linear regression modeling using stepwise entry criteria of p < 0.20 to identify factors potentially associated with antibody levels. Analyses were performed using SAS, version 9.4 (SAS Institute, Inc., Cary, NC). Methods and results are reported in accordance with Strengthening the Reporting of Observational Studies in Epidemiology (STROBE) statement^15^ and Standards for Quality Improvement Reporting Excellence (SQUIRE) guidelines (**Supplemental Table 1**).^16^

## RESULTS

### Population

Presented in **Table 1**, a total of 70 volunteers participated, age range was 62-97 years old with almost half in their 80s (49.3%) and the rest split between younger (22.5%) and older (28.2%). Two-thirds were female (60%) and almost all participants were white (97.1%). The frailty indices indicated moderate to high functioning.

**Table 1.**
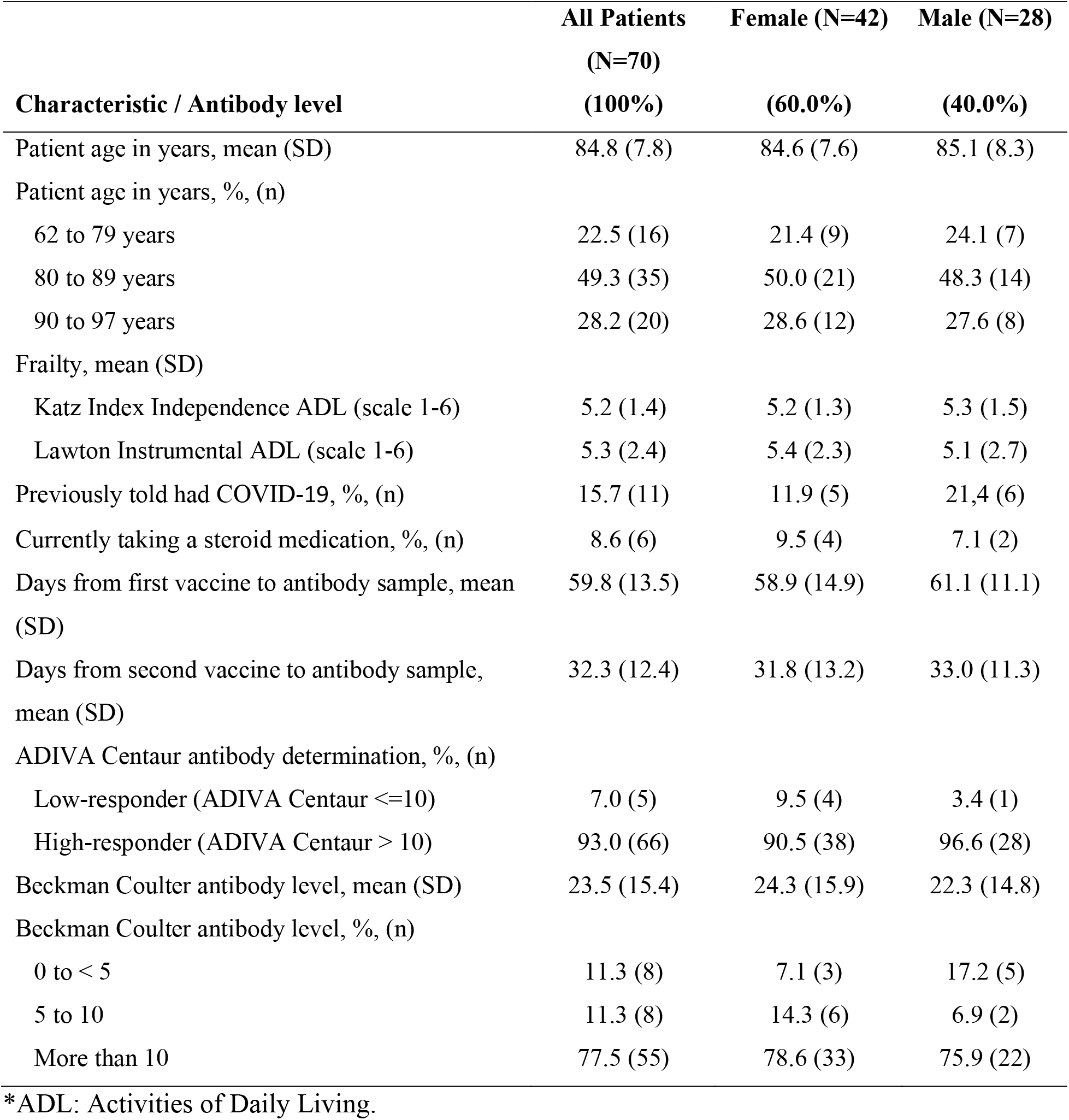
Descriptive Characteristics and Antibody Levels in Residents at Assisted Living, Personal Care, and Independent Living Facilities.

### Study Outcomes

All participants provided sera to be tested for antibodies to SARS-CoV-2, had undergone two rounds of vaccination (Moderna 98.6%, Pfizer 1.4%) within the prior 50 days, and one in six had recovered from COVID-19 infection (15.7%). Antibody levels were determined using two FDA Emergency Use assays. All participants were found to have antibodies to SARS-CoV-2; one was deemed non-reactive by the Beckman Coulter assay, having an extinction coefficient < 1, but was assessed as reactive by the more sensitive ADIVA Centaur assay.^13^ There is wide variation in relative levels of antibodies as determined by extinction coefficients. While the sample size is modest, which hinders statistical power, we found that antibody levels trended lower with male sex (standardized beta coefficient (β)=-0.11, p=.33) (**Figure 1**), advanced age (β=-0,18, p=.11), current use of steroids (β=-0,22, p=.07), and longer length of time from vaccination (β=-0.13, p=.28) (**Figure 2**). In participants who previously tested positive for COVID-19 (n = 11), antibody levels trended higher (β=0.18, p=.12), though one participant had very low levels of antibodies suggesting that prior infection does not guarantee a strong response.

**Figure 1.**
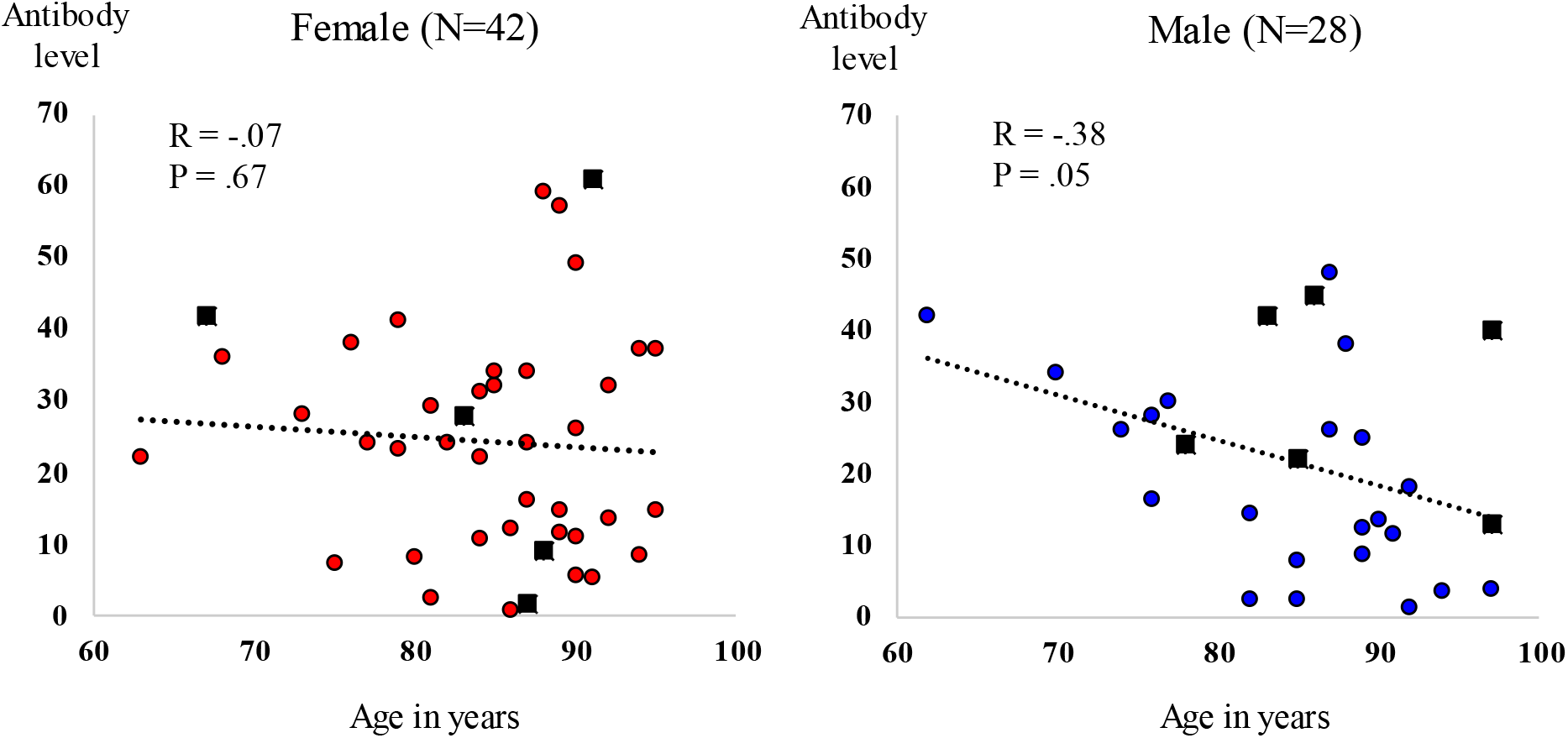
Scatter plot of patient age (x-axis) by Beckman Coulter antibody level (y-axis). Females (left plot) with red filled dots depicting participants without a prior history of COVID-19, and black filled rectangles depicting participants with a prior history of COVID-19. Males (right plot) with blue filled dots depicting participants without a prior history of COVID-19, and black filled rectangles depicting patients with a prior history of COVID-19.

**Figure 2.**
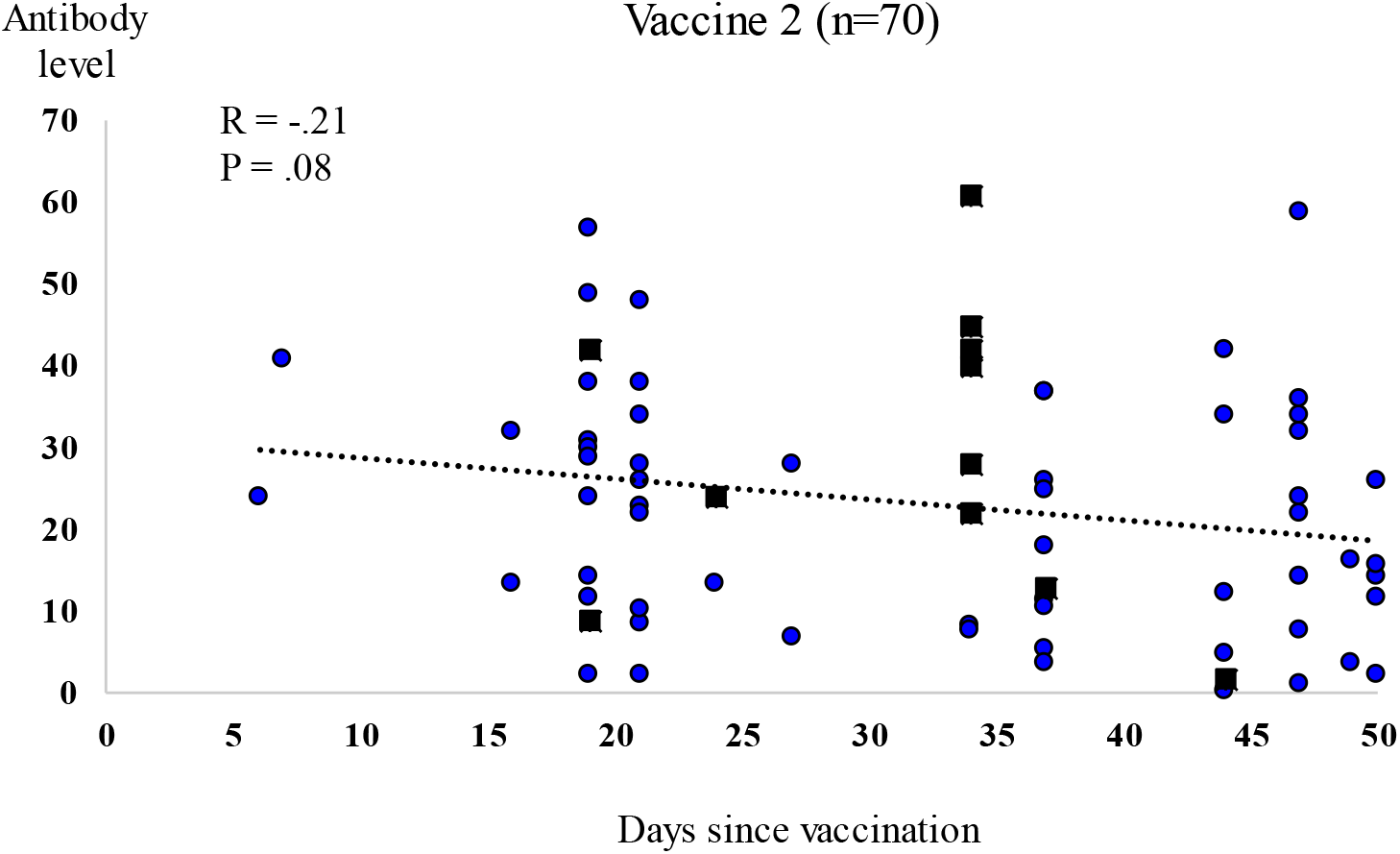
Scatter plot of days since second vaccination (x-axis) by Beckman Coulter antibody level (y-axis). Blue filled dots depict patients without a prior history of COVID-19, and black filled rectangles depict patients with a prior history of COVID-19.

## DISCUSSION

The results indicate that UPMC Senior Communities assisted living, personal care, and independent living residents did mount a detectable level of antibody responses – though antibody levels varied significantly among the individuals. Demonstration of vaccine response in this population, along with observational data demonstrating reductions of COVID-19 following implementation of vaccination,^2^ supports the argument for reopening facilities in the immediate period following vaccination.

This study has several limitations. Importantly, the presence of antibody levels does not necessarily confirm immunity. As with other viral infections, immunity to SARS-CoV-2 infection is complex and influenced by B and T cell responses and the innate immune system.^17^ Level of antibody, quality of antibodies produced, presence of neutralizing antibodies, and duration of antibody presence are all important unknowns in this population.^18^ Thus, the implications of these levels of antibodies in preventing COVID-19 disease must be determined by clinical follow-up, and incorporated into ongoing facility risk assessment as recommended.^4^ The modest sample limits the precision in estimates and conclusions drawn, particularly in stratified analyses. Participants were volunteers and likely to be healthier than non-participants, and individuals with known immunosuppression were excluded.

## CONCLUSIONS/IMPLICATIONS

Published recommendations regarding reopening of post-acute and long-term care settings focus on the process for reopening.^3-8^ Results from this study are part of an effort to assess population level of risk in reopening strategies for the residents at assisted living, personal care, and independent living facilities. The data reassures that moderate to higher functioning adults, even of advanced age, do mount detectable antibody responses when vaccinated with COVID-19 mRNA-based vaccines. These individuals demonstrate IgG within a range considered protective from other studies.^14^ This suggests that vaccination is functional and appropriate in these individuals. However, protective effects of such vaccination programs in advanced age residents at assisted living, personal care, and independent living facilities remain to be determined in larger studies.

## Data Availability

Raw data from this analysis is not permissible.

## Acknowledgments

The authors thank the UPMC COVID-19 Antibody Work Team including but not limited to Debbie Albin, Derek C. Angus, Brian Bachowski, J. Ryan Bariola, Robert Bart, Lorraine Brock, Christian Carmody, Suzanne Colilla, Jeanne Cunicelli, Maris Dauer, Jonna R. Drost, Utibe Ralph Essien, Ghady Haidar, David Huang, Allison Hydzik, Matthias Kleinz, Gloria A. Kreps, Joon Sup Lee, Kelsey Linstrum, Holly L. Lorenz, Genna M. Mancine, Oscar Marroquin, Susan C. Martin, Rhonda Martini, Erin McCreary, Thomas McGough, Jeffery C. McKibben, Bryan J. McVerry, Heather Mediate, Russell Meyers, Tami Minnier, Stephanie Montgomery, Stacy Parker, Jeffery Porter, Kevin Quinn, Mark Schmidhofer, Christopher Seymour, Judy Shovel, Eileen Simmons, Graham Snyder, Colleen Sullivan, Richard J. Wadas, Knox Walk, Dean E. Walters, Mary Wisniewski, Paul C. Wood, Tabitha Wybiral, Donald Yealy, Wendy L. Zellner, and their entire teams.

## Funding Statement

This work received no external funding. Internal funding from the UPMC Hospital System had no control over the study.

## Conflict of Interest Disclosure

None of the authors received any payments or influence from a third-party source for the work presented, and none report any potential conflicts of interest.

## SUPPLEMENTAL TABLE

**Table S1.** Checklist: Strengthening the Reporting of Observational Studies in Epidemiology (STROBE) and Standards for Quality Improvement Reporting Excellence (SQUIRE) 2.0 guidelines.

|  | Item No. | STROBE items | Location in manuscript where items are reported | SQUIRE items | Location in manuscript where items are reported |
| --- | --- | --- | --- | --- | --- |
| **Title and abstract** |  |  |  |  |  |
|  | 1 | (a) Indicate the study's design with a commonly used term in the title or the abstract (b) Provide in the abstract an informative and balanced summary of what was done and what was found | Abstract, Pages 2-3 | Title: Indicate that the article concerns an initiative to improve healthcare.<br>Abstract: This is a summary of your work and is the most important section to attract a reader's attention. Please ensure you include a brief background to the problem, the method for your quality improvement project, the overall results and conclusion. | Title Page 1<br>Abstract, Pages 2-3 |
| **Introduction** |  |  |  |  |  |
| Background rationale | 2 | Explain the scientific background and rationale for the investigation being reported | Introduction, Pages 4-5 | Background information about the problem and up-to-date, research and knowledge from the literature. | Introduction, Pages 4-5 |
| Objectives | 3 | State specific objectives, including any prespecified hypotheses | Introduction, Page 4-5 | Summarise your problem and the focus of your project. | Introduction, Page 4-5 |
| **Methods** |  |  |  |  |  |
| Study Design | 4 | Present key elements of study design early in the paper | Methods, Page 5 | Describe any reasons or assumptions that were used to develop the intervention(s) and reasons why you expected them to work. | Methods, Pages 5-6 |
| Setting | 5 | Describe the setting, locations, and relevant dates, including periods of recruitment, exposure, follow-up, and data collection | Methods, Page 5 |  |  |
| Participants | 6 | (a) Cohort study - Give the eligibility criteria, and the sources and methods of selection of | Methods, Page 5 |  |  |
|  |  | participants. Describe methods of follow-up<br>*Case-control study* - Give the eligibility criteria, and the sources and methods of case ascertainment and control selection. Give the rationale for the choice of cases and controls<br>*Cross-sectional study* - Give the eligibility criteria, and the sources and methods of selection of participants<br>*(b) Cohort study* - For matched studies, give matching criteria and number of exposed and unexposed<br>*Case-control study* - For matched studies, give matching criteria and the number of controls per case | Methods, Page 5 |  |  |
| Variables | 7 | Clearly define all outcomes, exposures, predictors, potential confounders, and effect modifiers. Give diagnostic criteria, if applicable. | Methods, Pages 5-6 | Explain your strategy for improvement and discuss how you implemented your study. | Methods, Pages 5-6 |
| Data sources/<br>measurement | 8 | For each variable of interest, give sources of data and details of methods of assessment (measurement). Describe comparability of assessment methods if there is more than one group | Methods, Pages 5-6 |  |  |
| Bias | 9 | Describe any efforts to address potential sources of bias | Methods, Pages 5-6 |  |  |
| Study size | 10 | Explain how the study size was arrived at | Methods, Pages 5-6 |  |  |
| Quantitative variables | 11 | Explain how quantitative variables were handled in the analyses. If applicable, describe which groupings were chosen, and why | Methods, Pages 5-6 |  |  |
| Statistical methods | 12 | (a) Describe all statistical methods, including those used to control for confounding<br>(b) Describe any methods used to examine subgroups and interactions<br>(c) Explain how missing data were addressed<br>(d) *Cohort study* - If applicable, explain how loss to follow-up was addressed<br>*Case-control study* - If applicable, explain how matching of cases and controls was addressed<br>*Cross-sectional study* - If applicable, describe analytical methods taking account of sampling strategy<br>(e) Describe any sensitivity analyses | Methods, Page 6 |  |  |
| Participants | 13 | (a) Report the numbers of individuals at each stage of the study (*e.g.*, numbers potentially eligible, examined for eligibility, confirmed eligible, included in the study, completing follow-up, and analysed)<br>(b) Give reasons for non-participation at each stage.<br>(c) Consider use of a flow diagram | Methods, Pages 6-7 |  |  |
| Descriptive data | 14 | (a) Give characteristics of study participants (*e.g.*, demographic, clinical, social) and information on exposures and potential confounders<br>(b) Indicate the number of participants with missing data for each variable of interest<br>(c) *Cohort study* - summarise follow-up time (e.g., average and total amount) | Results, pages 6-7 |  |  |
| Outcome data | 15 | *Cohort study* - Report numbers of outcome events or summary measures over time<br>*Case-control study* - Report numbers in each exposure category, or summary measures of exposure<br>*Cross-sectional study* - Report numbers of outcome events or summary measures | Results, Page 7 |  |  |
| Main results | 16 | (a) Give unadjusted estimates and, if applicable, confounder-adjusted estimates and their precision (e.g., 95% confidence interval). Make clear which confounders were adjusted for and why they were included<br>(b) Report category boundaries when continuous variables were categorized<br>(c) If relevant, consider translating estimates of relative risk into absolute risk for a meaningful time period | Results, Page 7 | Provide a summary of what your results showed. Comment on whether there were any unintended consequences such as unexpected benefits, problems, failures or costs associated with the intervention(s). | Results, pages 6-7 |
| Other analyses | 17 | Report other analyses done—e.g., analyses of subgroups and interactions, and sensitivity analyses | Results, Page 7 |  |  |
| **Discussion** |  |  |  |  |  |
| Key results | 18 | Summarise key results with reference to study objectives | Discussion, pages 7-8 | Comment on the strengths of the project. Describe any problems you faced and how you navigated these. | Discussion, pages 7-8 |
| Limitations | 19 | Discuss limitations of the study, taking into account sources of potential bias or imprecision. Discuss both direction and magnitude of any potential bias | Discussion, Page 8 | Reflect on your project's limitations. | Discussion, Page 8 |
| Interpretation | 20 | Give a cautious overall interpretation of results considering objectives, limitations, multiplicity of analyses, results from similar studies, and other relevant evidence | Discussion, Pages 7-8 | Describe whether chance, bias, or confounding have affected your results and whether there was any imprecision in the design or analysis of the project. Are more data points required? | Discussion, Pages 7-8 |
| Generalisability | 21 | Discuss the generalisability (external validity) of the study results | Discussion, Page 7-8 | Comment on the limits of generalisability. | Discussion, Page 7-8 |
| **Conclusions** |  |  |  |  |  |
|  |  |  |  | The point of the conclusion is not to rewrite the whole project, but to give an overview of how the whole project was conducted, what it achieved, and some personal reflections. | Conclusions Pages 8-9 |
| **Other Information** |  |  |  |  |  |
| Funding | 22 | Give the source of funding and the role of the funders for the present study and, if applicable, for the original study on which the present article is based | Page 8 |  |  |

## Notes

### Competing Interest Statement

The authors have declared no competing interest.

### Clinical Trial

This study was not a clinical trial.

### Author Declarations

This project underwent review and was granted ethical approval as a quality improvement study by the UPMC Quality Improvement Review Committee (Project ID: 3250), the ethics, regulatory, and legal oversight body for protecting patient/participant rights, confidentiality, consent (including waiver of consent), and the analysis and dissemination of deidentified data within the UPMC system.

